# EFFECTS OF LIFESTYLE ACTIVITY LEVEL ON ANTICIPATORY LOCOMOTOR ADJUSTMENTS FOR PEDESTRIAN CIRCUMVENTION

**DOI:** 10.1101/2024.09.10.24313424

**Authors:** Joris Boulo, Margaux Simon, Bradford J. McFadyen, Andreanne K. Blanchette

## Abstract

Navigating public environments requires adjustments to one’s walking patterns to avoid stationary and moving obstacles. It is known that physical inactivity induces alterations in motor capacities, but the impact of inactivity on anticipatory locomotor adjustments (ALA) has not been studied. The purpose of the present study was to compare ALAs and related muscle co-contraction during a pedestrian circumvention task between active (AA) and inactive young adults (IA). Thirteen AA and thirteen IA were placed in a virtual environment simulating a public park. Participants circumvented virtual pedestrians walking towards them. Walking speed, onset of deviation, clearance, foot placement strategies and muscle co-contraction were analysed. IA exhibited slower walking speeds compared to the AA during circumvention condition but not during unobstructed walking condition. The distance at the onset of trajectory deviation was larger for IA. Both groups increased co-contraction for pedestrian circumvention at the ankle and left hip and IA displayed greater ankle co-contraction overall. No significant group differences were observed in minimum clearance. This study suggests that an inactive lifestyle influences ALAs by inducing a cautious behavior during pedestrian circumvention.

## 1. Introduction

Navigating public environments is an essential aspect of everyday human activities. Avoiding both stationary (such as benches, poles, fire hydrants) and moving obstacles (such as pedestrians, animals) constitutes a significant part of living in the community (Musselman & Yang, 2007; Shumway-Cook et al., 2002). This locomotor navigation necessitates adaptations to walking pattern to avoid collision. Olivier et al. (2012) demonstrated that, in a perpendicular obstacle crossing setup, individuals initiated gait adjustments to prevent collisions as soon as they anticipated path intersections within less than one meter. In order to circumvent, pedestrians rely on visual cues from the person ahead of them. Then, the two pedestrians choose a collaborative circumvention strategy thereby preventing a collision (Bonsch et al., 2016; Olivier et al., 2013). In a frontal circumvention, anticipatory locomotor adjustments (ALA) are characterized by a simultaneous horizontal reorientation of the head and trunk (Vallis and McFadyen, 2003), following a lateral deviation of the center of mass (CoM) travel path (Fiset et al., 2020). Modifications in ALAs lead to an elliptical personal space, which allows individuals sufficient time to make further adjustments if necessary (Gérin-Lajoie et al., 2005).

To initiate the lateral deviation of the center of mass (CoM) required for circumvention, two lower limb movement strategies are possible: 1) side stepping (SS) where the inside leg (further from the obstacle) steps away from the outside leg to increase step width; and 2) cross stepping (CS) where the outside leg crosses over the inside leg resulting in a decrease in step width (Darekar et al., 2017; Paquette & Vallis, 2010). The SS strategy involves a higher peak CoM velocity and an increased stability but requires a significantly greater lateral impulse compared to the CS strategy (Acasio et al., 2017). The increased step width observed in SS suggests a more cautious strategy, as it provides a larger base of support (Paquette et al., 2008).

Previous studies have shown that the expression of ALAs depends on personal factors, such as age or physical skills. For example, it has been observed that older adults exhibit a relatively cautious approach in comparison to young adults by keeping a larger personal space (Gérin-Lajoie et al., 2006) and deviating farther from the obstacle (Rapos et al., 2019). It has also been shown that sport-specific skills influence obstacle avoidance abilities. For example, rugby players initiate an avoidance strategy significantly later than the non-athletes suggesting that sport-specific training may improve perception of one’s action capabilities, environmental perception and in turn their action skills (Fajen et al., 2008; Pfaff & Cinelli, 2018).

Sedentary and inactive lifestyles have been shown to significantly decrease muscular strength (Larsson et al., 1978; Thorstensson et al., 1977), reduce social interactions (Kraut et al., 1998), and increase social anxiety (Rubin et al., 2001). A model developed and latter confirmed by Stodden et al. (2008, 2009) hypothesized a strong relationship between motor skills and physical activity. Based on this model Carson Sackett & Edwards (2019) showed that increased physical activity (MET/week) was correlated with greater perceived self-competence and increased motor skill performance (object control, speed/agility, upper limb coordination). In the same vein, sedentary young adults have also been shown to have slower responses to perturbations compared to their active counterparts. Brinkerhoff et al. (2022) demonstrated that inactive adults responded less quickly to sudden perturbations in the speed of one of the belts of a split-belt treadmill. Finally, a study also revealed that sedentary individuals exhibit more co-contractions during unobstructed walking than active individuals (da Fonseca et al., 2006). This energetically inefficient contraction of antagonist muscles is often used in order to stabilize a joint (Winter, 2009). These findings indicates that a sedentary and inactive lifestyle may influence motor skills and abilities, potentially affecting community walking. However, the specific impact of inactivity on strategies employed during obstacle circumvention in daily navigation remain unclear.

The purpose of the present study was to compare the impact of physical activity level on ALAs including muscle activation during a daily pedestrian circumvention task in a virtual environment simulating a public park. We hypothesized that due to their reduced motor abilities, inactive individuals would exhibit more cautious ALAs that would likely lead to increased co-contractions and different foot placement strategies compared to an active group.

## 2. Materials and methods

### 2.1. Participants

Thirteen healthy active adults (AA; 7 females) and 13 healthy inactive adults (IA; 7 females) were recruited to participate in the study. Participants of the AA group had to be engaged in at least 3 sessions of high-intensity physical activity per week achieving a minimum of at least 1500 Metabolic Equivalent Task (MET) minutes/week (Forde, 2018). Participants of the IA group were required to engage in a sedentary behavior for more than 8 hours per day and not being engaged in any organized physical activity, according to the International Physical Activity Questionnaire (IPAQ; Craig et al., 2003; Gauthier et al., 2009). In order to minimize the influence of adipose tissue on electromyography (EMG) recordings, only participants with a body mass index (BMI) <30 were recruited (Bartuzi et al., 2010). Participants were excluded if they reported any musculoskeletal or neurological lesions affecting their gait, if they were unable to see and correctly describe the elements of the virtual environment (VE) (e.g., sign, door, bystanders) or if they reported a history of discomfort with screens or VR. The study was approved by the Research Ethics Board of the “Centre intégré universitaire de santé et de services sociaux de la Capitale-Nationale” and all participants provided informed written consent.

### 2.2. Experimental setup

A VE (Fig. 1) representing a 10 m by 10 m park was created in Unreal Engine (v.4.27; Epic Games, Cary, USA) and downloaded onto a head-mounted display (HDM; Meta Quest 2, 90 Hz, Menlo Pak, USA). The park has been designed to be the exact same size as the laboratory where the experiments were conducted. A soundtrack of bird sounds consistent with the park environment was added. Two virtual pedestrians representing a young man (shoulder width of 50 cm, 176 cm tall) and a young female (shoulder width of 40 cm, 166 cm tall) were created in MetaHuman. To control for differences due to the perceived gender of the virtual pedestrian in relation to one’s own gender identity, a picture of both virtual pedestrians walking was shown, and participants chose the virtual pedestrian they wished to interact with during the experiment.

**Fig. 1.**
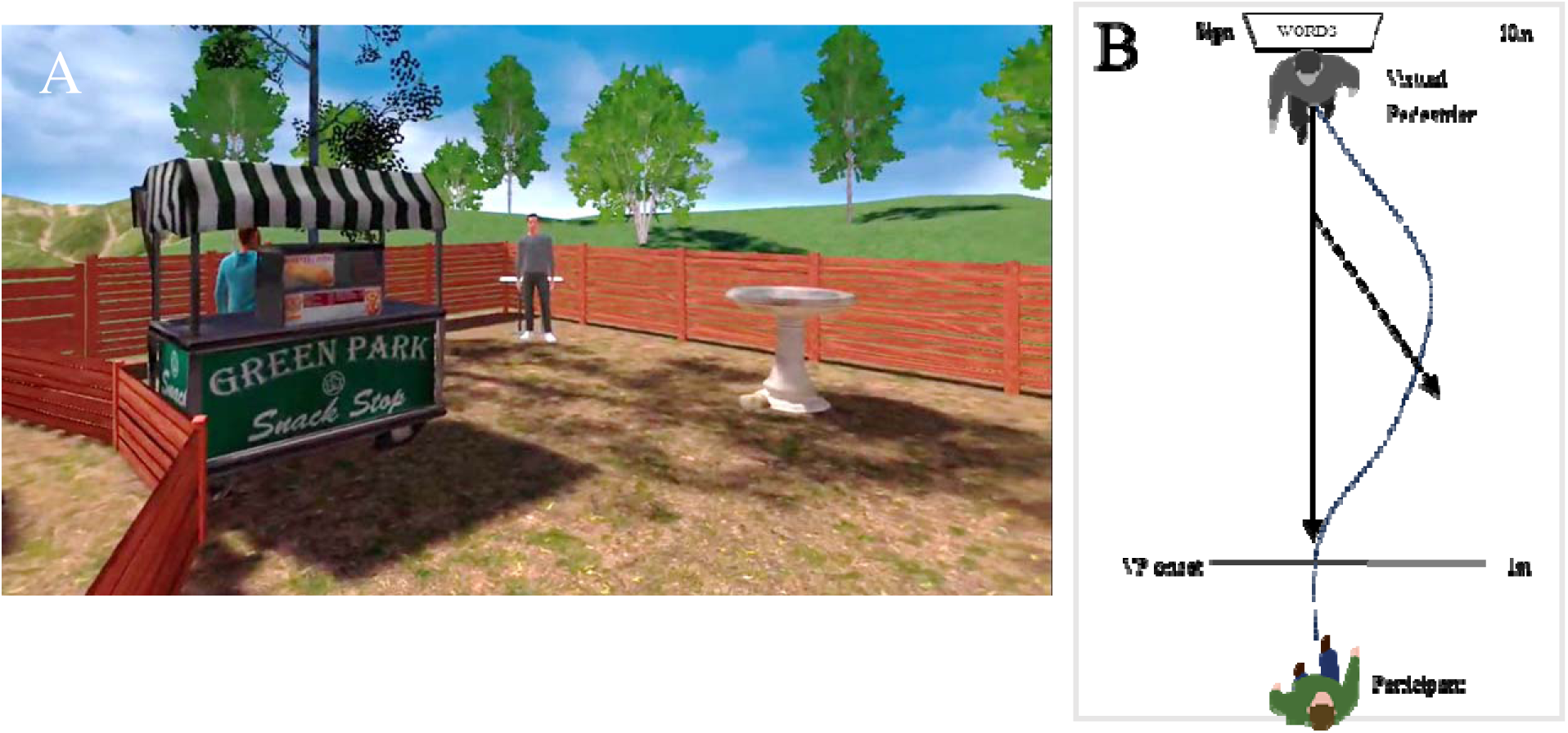
(A) Virtual environment (VE) as seen by the participant at the beginning of a trial; (B) representation of the VE seen from the top.

The psychometric qualities of the Meta Quest 2 have been previously tested (Boulo et al., 2023). The data from this study showed significant differences between the kinematic data from the Meta Quest 2 and the gold standard. Therefore, the acquisition of three-dimensional kinematic data were ensured by a 10-camera Vicon motion capture system (90 Hz; Oxford, United Kingdom). Four, non-collinear, reflective markers were affixed to the head-mounted display (HMD), trunk, pelvis and to both thighs, lower legs, and feet. To estimate the position of segment centers of mass (CoM), temporary markers were attached to anatomical landmarks including acromia, C7 and T10 vertebrae, sternal notch, xiphoid process, anterior-superior and posterior-superior iliac spine, medial and lateral malleoli, fifth metatarsal heads, heels, and toes during the calibration process.

Surface electromyography (EMG) was recorded (2000 Hz; Delsys, Natick, USA) with 8 sensors placed bilaterally on the gluteus medius (GM), hip adductors (ADD), lateral gastrocnemius (LG) and tibialis anterior (TA). It has been shown that these muscles are highly involved in unobstructed curve walking as well as directional changes (Choi et al., 2019; Courtine et al., 2006). Electrode placements were guided by the SENIAM (Surface ElectroMyoGraphy for the Non-Invasive Assessment of Muscles) recommendations (Hermens et al., 1999).

### 2.3. Protocol

In order to assess their activity level, participants completed the self-administered long version of the IPAQ (Gauthier et al., 2009) at the beginning of the experiment. To assess any differences between the two groups that might influence their navigation, the Liebowitz social anxiety scale was completed (Rubin et al., 2001; Yao et al., 1999) and leg dominance was identified (Gérin-Lajoie et al., 2005; Melick et al., 2017). Then, physical characteristics (height, mass) were measured, and visual abilities were tested (EDTRS score <0 logMAR to the Early Treatment Diabetic Retinopathy Study Letters (Kaiser, 2009).

To familiarize themselves with the HMD and the VE, participants were initially instructed to visually explore the VE with slow head turns. Subsequently, with assistance from the experimenter, they gradually transitioned to walking and exploring the environment. During the familiarization process, participants were guided by the experimenter to physically interact with the VE by touching a virtual wall corresponding to one within the lab, thereby enhancing their sense of presence in the VE. The familiarization process continued until they felt confident enough to navigate independently. Once the participant confirmed that they walked at their comfortable speed and that their walking speed remained consistent, their average baseline walking speed was measured across five trials. Each trial covered a distance of five meters within the VE. Virtual pedestrian walking speed was adjusted to 75% of this average walking speed.

For data collection, participants were instructed to start at a circular marker on the floor and then navigate within the park with the goal of reaching a sign positioned 10 m from the starting point. A simple word was written on this sign, which participants had to read out loud. Words were randomly selected from the French Manulex database for primary school level (Lété et al., 2004). This sign provided participants with a precise goal, yet their performance in word reading was not assessed. Three different conditions, described below, were proposed to the participants in a pseudo-random order (no more than 2 consecutive trials of the same condition). Different built environment elements were strategically positioned within the park to promote circumvention to the right (see Fig. 1).

An **Unobstructed walking condition** (no pedestrian) served as control trials for straight ahead walking. This condition was repeated 5 times. The circumvention condition was characterized by a virtual pedestrian walking directly toward the participant in a straight line (referred to as the **Virtual Pedestrian Strait condition** or VPS condition) forcing the participant to circumvent (10 trials). A **Catch condition** was added where the virtual pedestrian deviated from its regular path at a 25° angle toward the right and was repeated 5 times to make it difficult for the participant to anticipate the need for circumvention at the onset of each trial. In addition, the virtual pedestrian only initiated walking once the participant had progressed one meter. All the conditions where pseudo-randomised (no more than two consecutive trials of the same condition). If the participant tried to circumvent to the left, additional trials were performed until reaching 10 circumventions to their right side.

Following data collection, side effects induced by immersion, the sense of presence in the VE and perceived fatigue were documented using a French version of the Simulator Sickness Questionnaire (Kennedy et al., 1993) along with the Presence Questionnaire (Witmer & Singer, 1998) and a Borg CR10 scale (Borg, 1982; Williams, 2017), respectively.

### 2.4. Data analysis

Kinematic extracted from Vicon were low-pass filtered with a fourth order, zero-lag Butterworth filter at 9 Hz using MotionMonitor (Chicago, USA). Within the same software, three-dimensional head, and body center of mass (CoM) trajectories were calculated, as well as velocities. Kinematic data extracted from the HMD were imported into Matlab (USA) and low-pass filtered with a fourth order, zero-lag Butterworth filter at 9 Hz. Onset of trajectory deviation, defined as the last point where medio-lateral (ML) HMD velocity reached zero prior to lateral displacement exceeding two standard deviations of the average unobstructed walking condition trials, was identified for each trial of the VPS condition. Anterior distance from the VP at trajectory deviation onset was calculated as well as minimal clearance defined as the minimum radial distance occurring between the participant’s acromion and the VP’s shoulder up to the point of crossing. Median walking speed and maximal ML speed were calculated based on the strides between the onset deviation and the crossing for the VPS trials, or on 3 cycles in the middle of the scene for the unobstructed walking trials. To identify lower limb movement strategies while crossing, step width was measured by re-aligning the lab-based coordinates to be tangent to the participant’s path at the mid-point between two footsteps (Dingwell et al., 2023). A CS strategy was identified when the step width was negative indicating the outside leg crosses over the inside leg. Prevalence of CS was then reported as a measure of whether the participant used the CS strategy at least once between the onset of deviation and the crossing point for each trial.

EMG data were filtered with a fourth order, zero-lag Butterworth band-pass filter between 20 and 400 Hz (De Luca et al., 2010). The data were then rectified and smoothed using a moving average with a window length of 5% of the sampling frequency (100 points for a sampling frequency of 2 kHz) (Ives & Wigglesworth, 2003). Time normalization was performed by finding heel strikes and normalizing the cycle over 1000 points, i.e. between 2 consecutive heel strikes of the same leg. Amplitude normalization was applied using the mean peaks of 15 cycles, extracted from the unobstructed walking trials, for each participant and each muscle (Ghazwan et al., 2017). A co-contraction index (Fig. 2) was calculated for strides between the onset deviation and the crossing of the VP and calculated using the following equation (Winter, 2009):

**Fig. 2:**
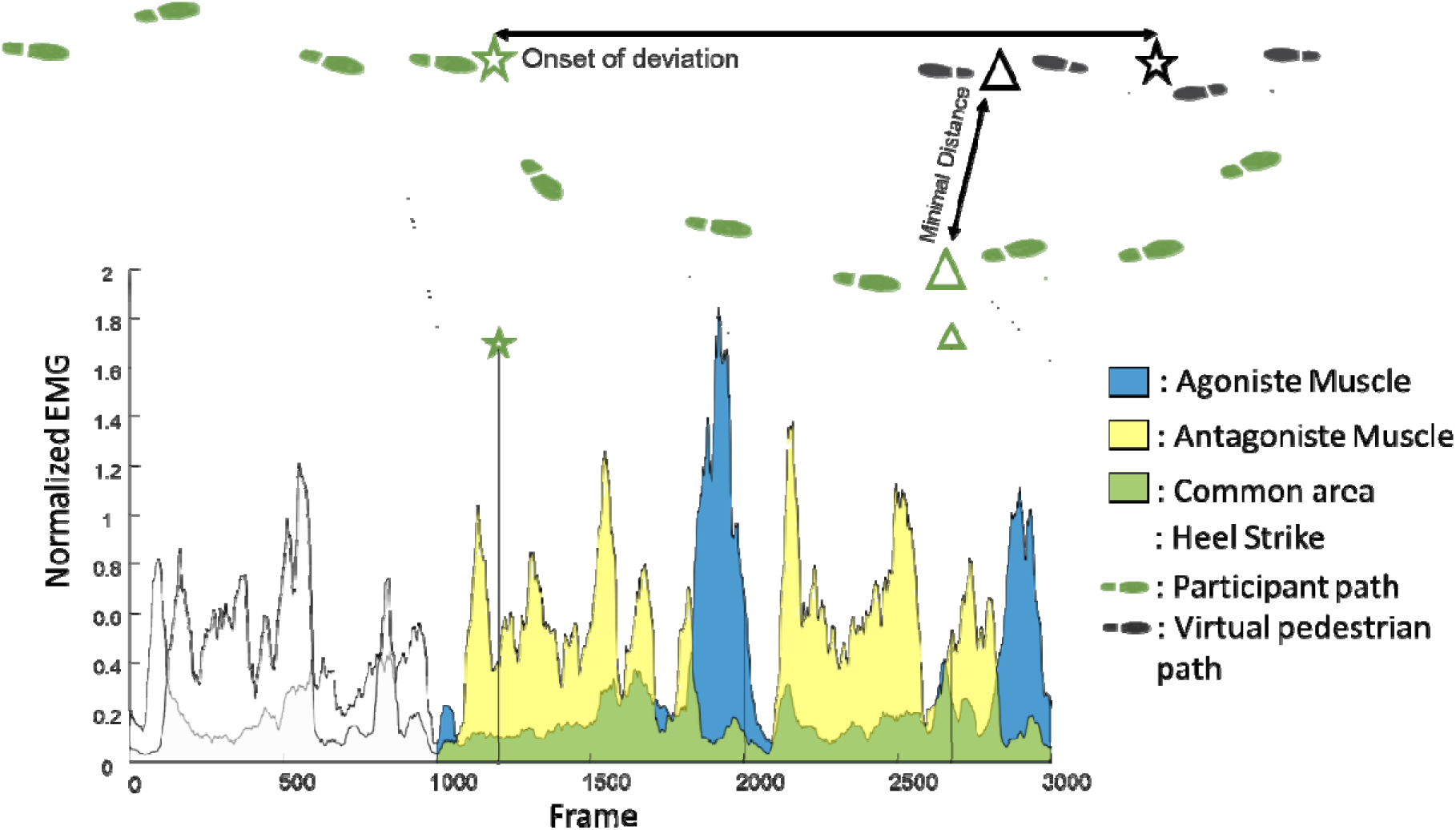
Representative example of the EMG activity profiles of TA and LG during a circumvention, with green area representing the co-contraction.

Within- and between-group comparisons of ALAs while circumventing a pedestrian were performed using nonparametric analysis of longitudinal data (NParLD). The latter was selected in order to be more conservative due to the small number of participants and to be less sensitive to outliers and issues of normality (Noguchi et al., 2012). A MixedNParLD model (F1-LD-F1 design) was used to compare dependent variables across groups (between factor) and repetitions of trials (within factor). For comparison between unobstructed walking and VPS condition, an additional dependant factor was added (F1-LD-F2) to compare speed and co-contraction between groups and between conditions. All analyses were conducted in Rstudio (Boston, USA) and Jamovi (Australia). The relative treatment effect (RTE) was reported for significant changes. Small, medium and large effects were respectively based on RTE threshold values of 0.56, 0.64 and 0.71 (Vargha & Delaney, 2000). Independent t-tests were used to compare group characteristics. To measure the prevalence of CS a generalized mixed model with repeated measures was performed for the prevalence of CS strategy.

## 3. Results

Except for their level of physical activity, the characteristics of participants in each group were similar (Table 1). There were no differences between the two groups in terms of age (*p* = 0.89), mass (*p* = 0.84), height (*p* = 0.11) and Body Mass Index (BMI; *p* = 0.31). As expected, there were significant differences between the two groups on the time spent doing moderate to high intensity sport (*p* < 0.01) and the time spent walking per week (p=0.01). The sports practiced by the AA were diverse, including triathlon, rowing, volleyball, rugby, and rock climbing. Although the AA group reported significantly lower involvement in sedentary behaviour during the week (*p* = 0.01), they still reported spending a median of 8 [6-9] h/day in sedentary positions. Both groups exhibited comparable scores on the Liebowitz Social Anxiety Scale (*p* = 0.69). All the participants of the IA group had a right dominance for footedness. Two participants of the AA group were left foot dominant. All but one participant chose to interact with the VP of the same gender. However, these between-group differences (footedness and the gender of the virtual agent with whom the participant interacts) did not show outlying results on the variables studied (i.e., they were within two standard deviations of the group mean). Finally, no collision with the virtual agent occurred for any participant.

**Table 1:** Characteristics of participants. Median and 25th-75th percentiles.

|  | Active Adults | Inactive Adults | *P*-Value |
| --- | --- | --- | --- |
| Age (y) | 25 [23-29] | 27 [22-31] | 0.89 |
| Mass (kg) | 64 [56-77.] | 62 [57-72] | 0.84 |
| Height (m) | 1.74 [1.68-1.79] | 1.67 [1.60-1.77] | 0.11 |
| Body Mass Index | 21.0 [20.2-22.9] | 23.1 [20.4-25.0] | 0.31 |
| Time spent doing moderate to high intensity physical activity per week (min) | 240 [210-450] | 0 [0-0] | $< \square .01$ |
| Time spent walking per week (min) | 330 [210-720] | 120[60-300] | **0.01** |
| Time spent in a sedentary behaviour during the week (h) | 8.00 [6.00-9.00] | 10.00 [8.00-10.05] | **0.01** |
| Time spent in a sedentary behaviour during the weekend (h) | 5.00 [3.00-7.00] | 8.00 [5.00-10.00] | 0.06 |
| Liebowitz Social Anxiety Scale | 20 [12-30] | 16 [13-42] | 0.69 |

No differences were found between the two groups on the Simulator Sickness Questionnaire (p=0.47) with scores of 0.13 [0.13-0.19] out of a total of four points (zero being no Sickness) for the AA and 0.13 [0.13-0.19] for the IA, suggesting a good tolerance of the VR. Participants of both groups showed similar (p=0.68) good immersion scores on the Presence Questionnaire in VR (AA: 6.20 [5.95-6.50]; IA: 6.20 [6.10-6.45]; out of seven points with seven being maximum presence) (Witmer & Singer, 1998). The protocol induced a very weak fatigue for either group based on the Borg test (AA: 0.00 [0.00-2.00]; IA: 1.00 [0.00-1.00]; p=0.72).

Regarding ALAs during circumvention, there was a between-group difference for walking velocity (*p* = 0.03; AA-RTE = 0.56; IA-RTE = 0.44), with the AA walking faster (median [25^th^ percentile – 75^th^ percentile]: 1.30 [1.22-1.44] m/s) than the IA (1.21 [1.11-1.24] m/s) (Fig. 3). However, this difference was not observed during unobstructed trials (*p* = 0.33). A group difference was also observed in maximal ML speed before crossing (*p* **<**□ 0.01; AA-RTE = 0.62; IA-RTE = 0.37), with the AA deviating at a faster speed (0.36 [0.25-0.43] m/s) compared to IA (0.24 [0.17-0.29] m/s).

**Fig. 3:**
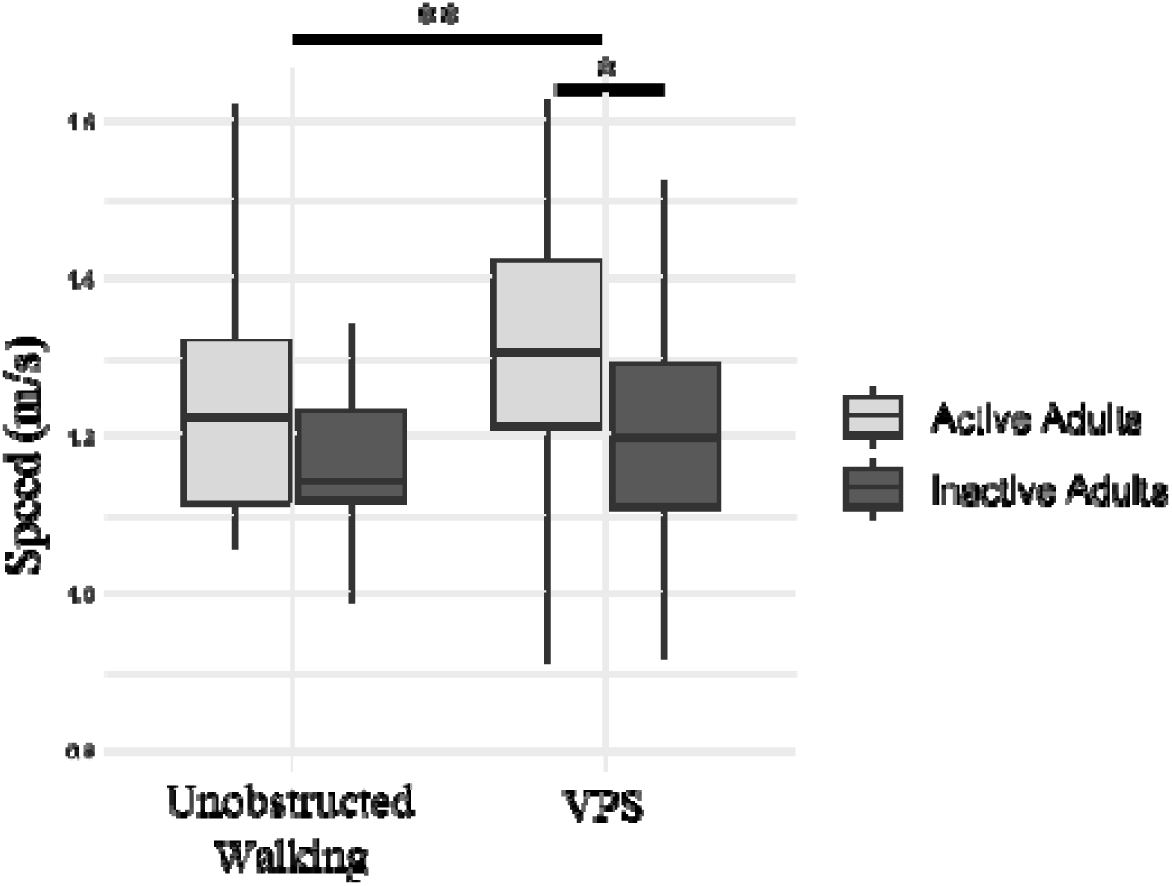
Walking speed (m/s) between active and inactive adults during unobstructed walking and Virtual pedestrian strait (VPS) conditions (Median, 25th and 75th percentiles, maximum, minimum); *: p<0.05, **: p<0.01

A statistically significant difference emerged between the two groups regarding the distance between the participant and the VP at the onset of deviation (*p***<**□0.01; AA-RTE = 0.41; IA-RTE = 0.59), with IA deviating farther from the VP (4.32 [3.74-4.68] m) compared to AA (3.19 [2.76-3.91] m). However, there was no significant difference in minimal clearance between groups (p = 0.42) (Fig. 4). During circumvention, AA tended to more frequently use a CS strategy compared to IA (*p* = 0.06). On average, participants in the AA group used a CS strategy in 56.94% (± 28.15) of the trials, while those of the IA group utilized it in 35.44 % (± 34.13) of trials (Shapiro-Wilk =0.93; Asymmetry coefficient = 0.041; Kurtosis = -1,24).

**Fig. 4:**
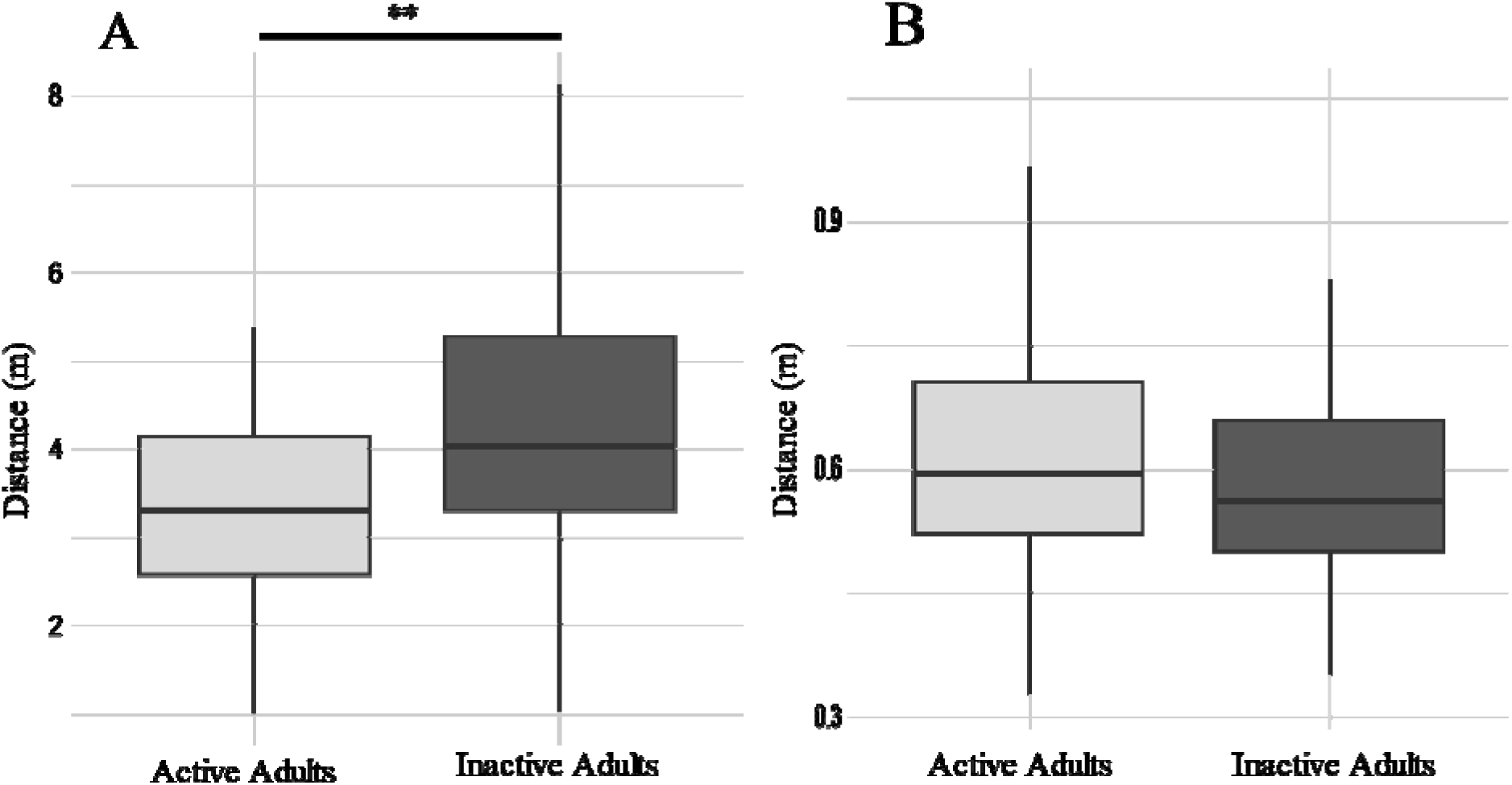
(A) Anteroposterior distance from the pedestrian at trajectory deviation onset and (B) minimal distance between the shoulders of the pedestrian and participant (clearance) at crossing point for active and inactive adults (Median, 25th and 75th percentiles, maximum, minimum); *: p<0.05, **: p<0.01

During circumvention, IA had greater co-contraction between TA and LG than AA on both sides. (Fig 5C-D: Right: *p* **<**□0.01; AA-RTE = 0.40; IA-RTE = 0.59; Left: p=0.002; AA-RTE = 0.39; IA-RTE = 0.60). In the same condition, no differences were noted in co-contraction between ADD and GM for either limb (Fig. 5A-B: Right: p = 0.96; Left: p = 0.19). In both groups, the circumvention around the pedestrian induced greater co-contraction between TA and LG compared to unobstructed walking on both sides (Fig. 5C-D: Right: p<0.01; AA-RTE = 0.40; IA-RTE = 0.60; Left: p<0.001; AA-RTE = 0.39; IA-RTE = 0.61). Additionally, the circumvention to the right elicited more co-contraction between ADD and GM on the left hip, although not on the right hip (Fig. 5A-B: Right: p=0.65; Left: p<0.01; AA-RTE = 0.47; IA-RTE = 0.52). During unobstructed walking condition, there were no between-group differences for co-contraction between ADD and GM for either limb (Fig. 5A-B). However, for this same condition, co-contraction between TA and LG was significantly still higher in the IA group compared to the AA group for both limbs (Fig. C-D: Right: p=0.016; AA-RTE = 0.37; IA-RTE = 0.62; Left: p<□0.01; AA-RTE = 0.35; IA-RTE = 0.64).

**Fig. 5:**
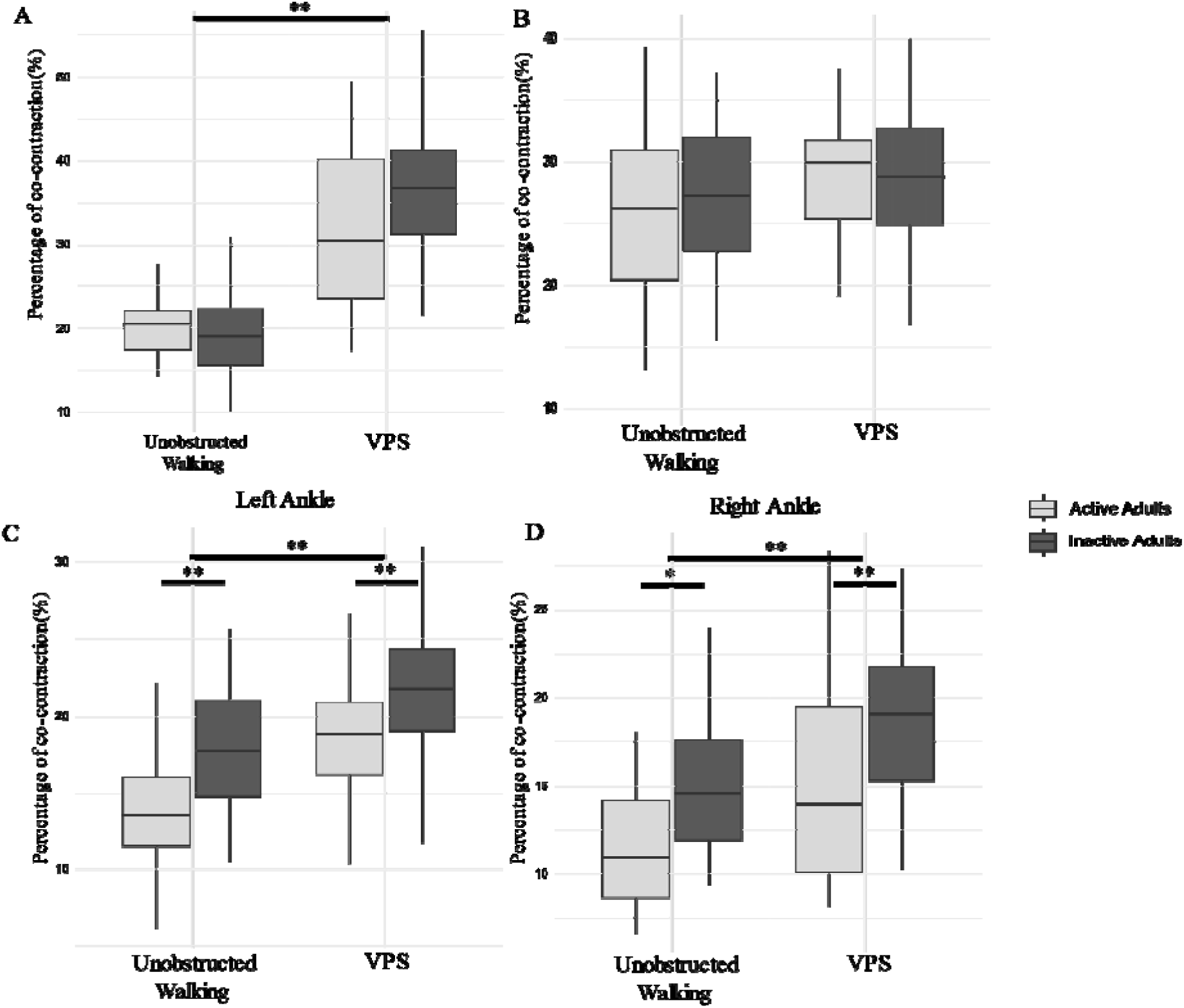
Active and inactive adults co-contraction between ADD and GM of the left hip (A) and the right hip (B), between TA and LG of the left ankle (C), and the right ankle (D) during unobstructed walking and virtual pedestrian strait (VPS) conditions (Median, 25th and 75th percentiles, maximum, minimum); *: p<0.05, **: p<0.01

## 4. Discussion

The purpose of the present study was to compare the impact of one’s level of daily activity on planning and executing pedestrian circumvention in relation to anticipatory locomotor adjustments (ALAs) and associated muscle co-contraction. We hypothesized that due to their reduced motor abilities, inactive individuals would exhibit more cautious ALAs during pedestrian circumvention than active adults. This hypothesis was confirmed as IA had a slower walking speed during circumvention and slower ML speed compared during circumvention. Moreover, the anteroposterior distance between the participant and the virtual pedestrian at trajectory deviation onset was greater for the IA compared to AA. However, no differences were observed in minimal clearance to the point of crossing between groups. As for the hypothesis about greater co-contractions, IA exhibited greater ankle co-contraction during the circumvention compared to AA. However, this greater ankle co-contraction was not specific to the circumvention condition, as it was also observed when IA walked in a straight line without obstacle circumvention.

These findings further support the role of activity level in influencing circumvention strategies. The finding that IA walked slower during circumvention compared to AA is consistent with the results reported by Niang & McFadyen (2005) who contrasted ALAs for stepping over an obstacle in a real-world environment between active and inactive young adults. The authors also noted a between-group difference during unobstructed walking. In the present study, both groups exhibited similar unobstructed walking capabilities, while circumvention abilities were influenced by their physical activity habits. For a circumvention task, an earlier deviation onset and a greater minimal clearance can be interpreted as a more conservative strategy (Gérin-Lajoie et al., 2005). The larger anteroposterior distance with a VP at the onset of deviation, as observed in the present study, would provide IA more time to perceive the situation and make necessary gait adjustments. The present study also revealed that IA tended to prefer the SS strategy over the CS strategy for foot placement. The SS strategy further supports a more cautious behaviour, as noted by Paquette et al. (2008). In a study comparing ALAs between rugby players and non-athletes, Pfaff and Cinelli (2018) demonstrated that athletes achieved higher maximal ML speed and deviated closer to the pedestrian compared to non-athletes. However, in this study the fact that the AA practiced different sports suggests that the observed differences between active and inactive are not related to a sport-specific training. Thus, while it is difficult to say with certainty whether the non-athletic group in Pfaff and Cinelli were more prudent, it does appear from the current study that the inactive, sedentary group practices more caution based on slowing of gait, further deviations, and safer foot placement strategies.

The current study showed that inactivity and sedentary behaviors result in increased co-contraction between TA and LG suggesting the need to increase joint stiffness at the ankle during walking. Co-contraction may lead to a diminished net effect of the agonist (Winter, 2009) and leads to heightened energy expenditure (Peterson & Martin, 2010). Di Nardo et al. (2015) also showed that co-contractions of TA and LG occur during normal walking to recover balance, control ankle stability, and manage knee flexion. Thus, joint stiffening is often characterized as a postural control method associated with cautious behaviour (Lee et al., 2017).

This study also showed that circumvention to the right induced greater co-contraction in the left hip muscles, regardless of the physical activity habits. This increased co-contraction may be related to the utilization of the CS strategy, which requires precise mediolateral foot placement. Co-contraction has been shown to play an important role when increased accuracy is needed (Gribble et al., 2003). We therefore hypothesize that deviating with a CS strategy represents a multi-joint challenge that requires an increased co-contraction to ensure a safe walk and avoid tripping. It cannot be ruled out that that right/left hip differences may have been influenced by the fact that EMG electrodes were placed lower than the SENIAM recommendations on the left limb and higher on the right limb. This decision was made to prevent the sensors placed on the muscle group of each respective lower limb from colliding during walking and avoid interfering with the natural gait of the participants.

Various factors might influence the IA to adopt more cautious behaviours in their ALAs. First, it has been shown that an inactive and sedentary lifestyle induces a poorer balance control in quiet standing (Minino, 2022; Zhu et al., 2021). Moreover, inactive behaviors are associated with significant declines in muscular strength (Larsson et al., 1978; Thorstensson et al., 1977), and proprioceptive abilities (Borges et al., 2018). Li et al. (2020) investigated the relationships between individual motor coordination, perceptions of physical abilities, and physical activity among a group of young adults aged between 17 and 23 years old. A strong correlation between perceived physical abilities and level of physical activity was found. Taken together, the decrease of neuromotor capabilities among inactive individuals may lead to lower confidence in their abilities to change direction compared to active individuals. This study suggests that premorbid physical activity level should be investigated when accessing locomotor navigation as individuals do not share the same baseline abilities.

This study had certain limitations. First, virtual reality (VR) tends to encourage a more cautious approach to avoidance behavior (slower walking speed, bigger clearance, bigger maximum deviation), as demonstrated in previous research (Berton et al., 2019; Bühler & Lamontagne, 2019). Additionally, it is important to recognize that, in addition to the basic visual feedback of VR, this experiment provided only auditory feedback related to collision. The incorporation of haptic feedback during pedestrian collisions could have enhanced the sense of presence and potentially affect behavioral responses (Berton et al., 2022). However, participants were provided with haptic feedback of the built environment during familiarization by touching a virtual wall that matched a real wall.

## 5. Conclusion

Physical inactivity and sedentary behaviour among young adults affect ALAs during pedestrian circumvention leading to more cautious path planning and execution of lateral deviations. Thus, physical inactivity may have an impact on daily locomotor navigation for healthy young adults. This cautiousness might result in decreased community walking and thus in more deconditioning. Moreover, this study suggests that prior level of physical activity should be taken into consideration when assessing locomotor navigation abilities of elderly individuals or those with neurological disorders or lesions such as an acquired brain injury. Finally, the changes caused by inactivity in such a common daily task highlight the need for increased emphasis on promoting physical activity.

## Data Availability

All data produced in the present study are available upon reasonable request to the authors

## Declarations

### Conflict of interest

The authors declare that they have no conflict of interest.

### Fundings

This project was funded by Natural Sciences and Engineering Research Council of Canada (RGPIN/191782-2023; BJM) and the Réseau provincial de recherche en adaptation-réadaptation (REPAR; 0101674). JB received a scholarship from the Centre interdisciplinaire de recherche en réadaptation et intégration sociale (Cirris).

## Acknowledgement

We would like to thank Jonathan Caron-Roberge and Félix Fiset for their valuable contribution, respectively in programming and technical assistance. We would also like to thank Mr. Gerard Mulvany and Prof. Stefan Greuter for sharing 3D printing plans for the plastic support used for the calibration frame.

## Author contributions

All authors contributed to the experimental design and project conceptualization. Data collection and management was completed by JB and MS. JB and MS conducted data processing and analyses, supervised by AKB and BJM. Figures and tables were created by JB. Statistical analyses were completed by JB. All authors contributed to the interpretation of the results. JB drafted the initial version of the manuscript while BJM, MS, and AKB conducted multiple revisions. All authors approved the final version.

## Notes

### Competing Interest Statement

The authors have declared no competing interest.

### Funding Statement

This project was funded by Natural Sciences and Engineering Research Council of Canada (RGPIN/191782-2023; BJM) and the Reseau provincial de recherche en adaptation-readaptation (REPAR; 0101674). JB received a scholarship from the Centre interdisciplinaire de recherche en readaptation et integration sociale (Cirris).

### Author Declarations

The study was approved by the Research Ethics Board of the “Centre intégré universitaire de santé et de services sociaux de la Capitale-Nationale” and all participants provided informed written consent.

